# Peripheral Biomarkers of Tobacco-Use Disorder: A Systematic Review

**DOI:** 10.1101/19004150

**Authors:** Dwight F. Newton

**Affiliations:** University of Toronto, Department of Pharmacology & Toxicology, Toronto, Ontario, Canada

**Author notes:** Corresponding Author Dwight Newton, Department of Pharmacology and Toxicology, University of Toronto, Toronto, Ontario, Canada.

**Keywords:** Tobacco-use disorder, nicotine dependence, biomarkers, HPA axis, cardiovascular disease, oxidative stress

## Abstract

**Introduction:** Tobacco use disorder (TUD) is a major worldwide healthcare burden resulting in 7 million deaths annually. TUD has few approved cessation aids, all of which are associated a high rate of relapse within one year. Biomarkers of TUD severity, treatment response, and risk of relapse have high potential clinical utility to identify ideal responders and guide additional treatment resources.

**Methods:** A MEDLINE search was performed using the terms *biomarkers, dihydroxyacetone phosphate, bilirubin, inositol, cotinine, adrenocorticotropic hormone, cortisol, pituitary-adrenal system, homovanillic acid, dopamine, pro-opiomelanocortin, lipids, lipid metabolism* all cross-referenced with *tobacco-use disorder*.

**Results:** The search yielded 424 results, of which 57 met inclusion criteria. The most commonly studied biomarkers were those related to nicotine metabolism, the hypothalamic-pituitary-adrenal (HPA) axis, and cardiovascular (CVD) risk. Nicotine metabolism was most associated with severity of dependence and treatment response, where as HPA axis and CVD markers showed less robust associations with dependence and relapse risk.

**Conclusions:** Nicotine-metabolite ratio, cortisol, and atherogenicity markers appear to be the most promising lead biomarkers for further investigation, though the body of literature is still preliminary. Longitudinal, repeated-measures studies are required to determine the directionality of the observed associations and determine true predictive power of these biomarkers. Future studies should also endeavour to study populations with comorbid psychiatric disorders to determine differences in utility of certain biomarkers.

## Introduction

Tobacco use remains one of the largest worldwide healthcare burdens despite decades of research and public awareness of adverse health outcomes, being responsible for over 7 million deaths annually (1). Addiction to tobacco products, tobacco use disorder (TUD) in DSM 5, is characterized by compulsive drug seeking and use, which continues despite harmful consequences to the individual (2). It is estimated that 60-80% of current daily cigarette smokers meet DSM 5 criteria for TUD, representing a substantial and largely unmet treatment challenge (3). Unfortunately, despite the development of cessation aids, 80-95% of patients attempting to quit will relapse within 1 year (4). TUD is complicated by high rates of comorbid psychiatric disorders with symptoms which overlap with those of TUD, creating diagnostic and treatment challenges (5). These realities highlight the importance of improved diagnostic and prognostic (both course of illness and treatment response) tools.

Biological markers of TUD hold great promise to address these problems which are pervasive across psychiatric disorders. Current diagnostic measures are based on identifying behavioural symptoms (e.g. loss of control, craving) and essentially no objective biologically-informed treatment selection tools exist (6). However, it is well understood that the underlying biology of TUD involves neuroplastic changes in reward circuitry occurring with repeated drug use and the formation of drug-salient environmental and interoceptive cues (7). Maladaptations of the dopamine (DA) system of the nucleus accumbens and ventral tegmental area occurring in addictive disorders have been extensively characterized, and drugs of abuse almost invariably induce changes in this circuitry with repeated use (7).

Neuroimaging has provided a window to study these structural and functional changes in humans, and identify biomarkers related to specific symptom dimensions, such as craving or impulse control, and clinical phenomena such as relapse and treatment response (8-10). Despite their immediate relevance to the pathophysiology of TUD, central nervous system (CNS)-based biomarkers are particularly costly to acquire and require specialized facilities and technical staff, greatly limiting their scalability and utility as predictive biomarkers. Such considerations are especially relevant when biomarkers are measured repeatedly over time, in the case of markers of treatment efficacy or relapse or in healthcare settings without neuroimaging equipment. Peripheral (non-CNS) markers, though potentially lacking direct representations of CNS pathophysiology, have the advantages of being easily collected, inexpensive, and can be rapidly integrated into ongoing clinical practice.

Recent reviews of biomarkers in addictive disorders have largely focused neuroimaging markers, which is warranted given the known similarity of neurobiological changes occurring in the mesolimbic DA system across drugs of abuse (8, 11, 12). Reviews of tobacco-using populations specifically have been limited by either focusing on all smoking populations without consistently quantified levels of dependence or examining a single class of biomarkers (e.g. nicotine metabolites) (13-16). Genetic markers, while clearly relevant to TUD clinical outcomes (17-19), have limited a ability to track dynamic changes over time. Dynamic, non-genetic, biomarkers are better suited to capturing features which change over time such as the course of treatment response or risk of relapse. A focus on TUD may be more clinically relevant than examining smokers regardless of dependence and tobacco use intensity. Though nicotine induces changes in nicotinic receptor expression after a single dose which persists after clearance of the drug, the neurobiological changes occurring in addiction involve systems beyond the specific pharmacology of the drug of abuse (20). Furthermore, a focus on TUD may capture a more severe subgroup of the tobacco-using population which may decrease the potential noise in biomarker signals occurring in less dependent groups.

Existing biomarkers for TUD, and SUDs more broadly, have generally been those related to drug exposure (6). In the case of nicotine, these have been expired carbon monoxide (CO) and cotinine measured in saliva, urine, or blood (6). Expired CO is increased acutely (6-9 hours) after combustible tobacco use, whereas cotinine, the primary metabolite of nicotine, has a longer half life (5-7 days) thus giving a longer-term measure of exposure to nicotine, regardless of the route of administration (21). However, growing evidence suggests that these, other exposure markers, are associated with dependence severity and risk of relapse (22, 23). Biomarkers not directly related to nicotine metabolism have been increasingly studied in TUD, such as markers of cardiovascular disease (CVD), inflammation, oxidative stress, and hypothalamic-pituitary-adrenal (HPA) axis reactivity (24-26). Such markers may be related to the underlying pathophysiology of TUD or relevant comorbidities, and thus may relate more strongly or specifically to symptom dimensions or relevant clinical outcomes.

Given the growth of the literature regarding biomarkers related to smoking and tobacco use, and the broader, or distinct, scope of existing reviews, a systematic review of the extant literature of peripheral biomarkers related to TUD will be performed. This review will focus on TUD and nicotine dependence specifically and aims to determine the peripheral biomarkers related to TUD and relevant clinical outcomes. Suggestions regarding future research directions and clinical implications will also be presented.

## Results

### Search Results and Study Selection

The results of the search yielded 424 studies in total, of which 57 met inclusion criteria. Of the 367 studies excluded (Figure 1): 142 were reviews or other non-original research articles, 90 did not measure a peripheral biomarker, 90 did not include TUD populations or measures of dependence, 40 reported biomarkers which were solely genetic, and 5 were not conducted in humans. An overview of the included studies can be found in supplementary table S1. In the following summary, the term *significant* will be omitted for ease of reading and any mention of increase, decrease, or correlation will be statistically significant (p<0.05).

**Figure 1.**
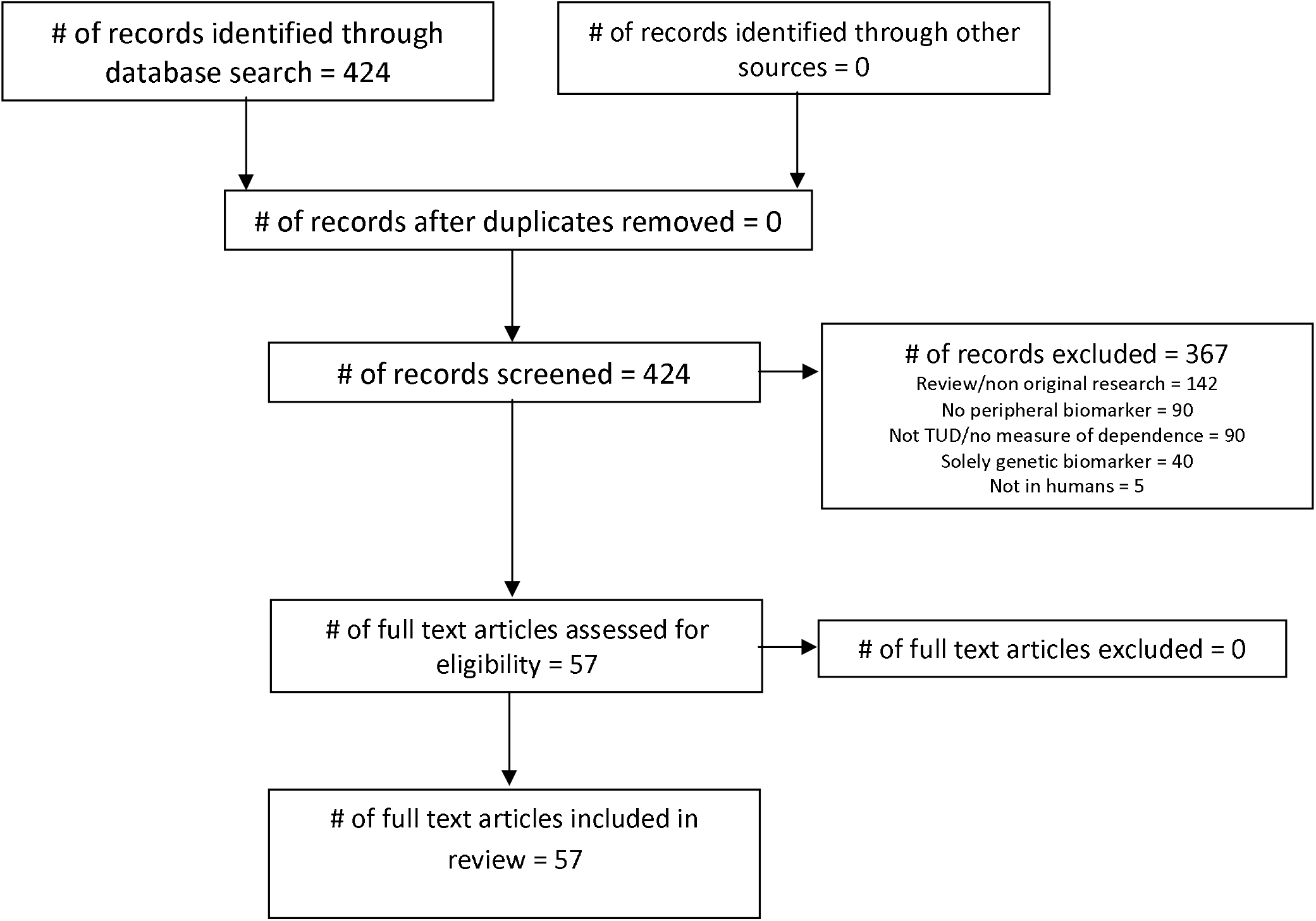
PRISMA diagram detailing the literature search workflow and inclusion and exclusion of potential studies.

### Markers of Exposure

Unsurprisingly, markers of exposure (also referred to as recency-of-use markers) are directly related to exposure to nicotine or toxicants present in cigarettes and other tobacco products. Those reported in the following studies include nicotine, cotinine, 3-hydroxycotinine (3-HC), carboxyhemoglobin (CHb: CO-bound hemoglobin), total nicotine equivalents (an omnibus measure of nicotine and its metabolites), carcinogenic tobacco-derived nitrosamines (NNK, NNAL, and NNN), polycyclic aromatic hydrocarbons (PAH) and alkaloids found in tobacco smoke. The nicotine metabolite ratio (NMR) is a commonly reported biomarker calculated from the ratio of cotinine to 3-HC, which also reflects CYP2A6 activity – the enzyme responsible for both nicotine to cotinine and cotinine to 3-HC metabolism (28). All studies below measure dependence using the Fagerström Test for Nicotine Dependence (FTND), or the Heaviness of Smoking Index (HSI) for pregnant populations.

A cross-sectional study of 239 smokers measuring urinary nicotine, cotinine, and 3-HC found that levels of all three of these markers were positively correlated with FTND scores (29). The *time to first cigarette* (TTFC) item of the FTND was positively correlated with urinary levels of nicotine, cotinine, and 3-HC. However, cotinine and 3-HC were not associated with cigarettes per day (CPD). These results were replicated in a separate study of 196 smokers which measured salivary cotinine only (30). Cotinine was associated with FTND scores, and the FTND item showing the strongest individual association with cotinine was again the TTFC. Urinary cotinine was also correlated with overall FTND scores in a study of 381 smokers (31). In this study a receiver operating characteristic (ROC) analysis indicated that urinary cotinine could differentiate highly dependent (FTND≥6) smokers from others with 71% sensitivity and 74% specificity. A large-scale study of 3585 smokers examining urinary nicotine equivalents and serum levels of cotinine and carboxyhemoglobin found that levels of each biomarker were associated with FTND scores, with TTFC showing the strongest correlation of all items (32). In contrast to the above studies, exposure markers were significantly associated with CPD. In a study of 204 smokers (69 black and 135 white), urinary nicotine equivalents, NNAL and PAH were measured (33). Surprisingly, nicotine equivalents and NNAL levels were correlated with greater FTND scores in white smokers, but this was not the case for black smokers. This demonstrates the need to continue developing psychometric tools for assessing nicotine dependence, as they may not be equally accurate across populations. A study measuring salivary nicotine and cotinine in 282 male smokers found that cotinine was significantly associated with FTND, but not nicotine, once again indicating the importance of measuring metabolites (34). These findings demonstrate that nicotine metabolites, especially cotinine, are related to severity of nicotine dependence and specific smoking features, such as the TTFC. Surprisingly, correlations between such recency-of-use markers and measures of intake alone (i.e. CPD) were less consistent.

Longitudinal studies of exposure markers have primarily been secondary analyses of randomized controlled clinical trials (RCTs) or non-randomized studies of smoking cessation, and thus offer an opportunity to look at potential predictive relationships with treatment response. One such secondary analysis of a 8-week RCT of nicotine replacement therapy (NRT), in the form of a 21mg/day transdermal nicotine patch, measured the impact of NMR at baseline on treatment response in 474 smokers (22). Those with a lower NMR (i.e. faster metabolizers) had a decreased likelihood of abstinence at week 8. This finding was replicated in a 8-week RCT of combined NRT (patch) and behavioural counselling which measured salivary NMR in 499 treatment-seeking smokers. It was found that patients with faster NMRs showed lower quit rates at 8-weeks (35). In the same study, NMR was not associated with any changes in craving or withdrawal symptoms. A third 8-week clinical trial of NRT (patch) measured plasma cotinine at baseline and after 1-week in 56 smokers. Percent cotinine replacement did not correlate with any smoking outcome.

Interestingly, this relationship between NMR and likelihood of cessation is also seen in short-term paradigms. A study using a 5-day contingency-management paradigm, in which smokers were monetarily rewarded for a negative expired CO reading on each of the 5 days, measured salivary cotinine in 103 non-treatment seeking smokers at intake (36). In this study, cotinine levels were predictive of expired CO levels at all timepoints, poorer study retention, lower chance of 100% abstinence, and shorter time to first positive CO reading. These studies indicate that nicotine metabolites, NMR especially, may be clinically salient biomarkers and individuals with fast NMRs may require further cessation aids. These findings also suggest an importance for composite biomarkers (e.g. NMR), which seem to relate more robustly to NRT response than cotinine alone. Indeed, NMR is likely particularly relevant to NRT given the fact that nicotine itself is the primary therapeutic agent, more so than non-NRT pharmacotherapy such as varenicline or bupropion.

Though the association between exposure markers and dependence established in cross-sectional studies is compelling, it cannot be determined from such studies if this relationship is modifiable. Studies which experimentally modulate nicotine exposure can, however. A secondary analysis of a 6-week RCT investigating very low nicotine content cigarettes (VLNCs) (0.4-2.4 mg/g nicotine) measured salivary NMR and urinary NNAL at intake, 2-weeks, and 6-weeks (37). Subjects randomized to VLNCs had lower NNAL and FTND scores after 6-weeks. These findings were broadly replicated in another study of VLNCs, in which 20 non-treatment seeking smokers were progressively tapered to a 0.1mg nicotine VLNC over 6-weeks (38). Subjects showed reduced FTND scores, cotinine, cHb, and NNAL at 6-weeks compared to baseline, despite the presence of compensatory smoking. cHb and NNAL, but not cotinine, remained decreased at 10-weeks. A similar approach was used in a 6-week semi-blinded RCT of VLNCS in which subjects were randomized to either 0.05mg nicotine VLNC (n=53), 0.3mg VLNC (n=52), or a 4mg nicotine lozenge (n=60) in which urinary cotinine, NNAL, NNK, NNN, and PAH were measured at 2 and 6-weeks (39). Patients receiving 0.05mg VLNCs or lozenges showed reduced nitrosamines levels at 6-weeks compared to the 0.3mg group, and the 0.05mg VLNC group had lower cotinine at 2 and 6-weeks compared to both groups. Though this last study not directly measure dependence, these findings show that reducing tobacco (and nicotine) intake via VLNCs can reduce markers of nicotine metabolites, carcinogenic tobacco-derived nitrosamines, and severity of nicotine dependence. Though it is unsurprising that nitrosamines and other nicotine metabolites are decreased after VLNC substitution, it is notable that their decrease is correlated to, or coincides with, decreased dependence. Lastly, though the above studies were performed in otherwise smoking populations, and these markers maintain correlations with dependence in smokeless tobacco users (40, 41), and pregnant smokers (42).

### Stress Response

The HPA axis is a key mediator of the physiological stress response, originating with hypothalamic release of corticotropin-releasing hormone (CRH), stimulating the anterior pituitary to release adrenocorticotropic hormone (ACTH) into the circulation (43). ACTH, in turn, stimulates the production and release of cortisol from the adrenal glands, thus ACTH and cortisol are commonly used peripheral measures of HPA axis function. Non-HPA axis stress biomarkers have also been investigated in the context of TUD, including alpha-amylase, prolactin, and catecholamines such as epinephrine and norepinephrine. Alpha-amylase is released in saliva during stress and sympathetic nervous system activity (44, 45) and prolactin is elevated after stress in a diffuse manner across the body (46). Actively smoking is known to induce an acute biological stress response and increases circulating cortisol levels (47). Both steady-state and stress-induced levels of HPA-axis markers have been investigated as biomarkers in TUD populations.

Most studies measuring biomarkers of stress have used psychosocial or pharmacological paradigms to induce stress responses, and then determine the relationships between stress hormone levels and clinical outcomes. The Trier Social Stress Test (TSST) has been widely used to induce stress, which involves a public speaking task and surprise mental arithmetic task performed for an unsupportive audience (48). A study of 64 smokers measured salivary cortisol and alpha-amylase before, and during the response to, the TSST (49). Those with low nicotine dependence (median split of FTND scores) showed similar baseline cortisol levels to those with high dependence, but showed a greater increase in cortisol in response to the TSST. Highly dependent smokers showed a blunted cortisol response, i.e. no increase after the TSST. This blunted cortisol response has been further replicated in 23 dependent smokers (50), 23 non-dependent smokers, and 25 controls (salivary cortisol), 116 smokers and 38 controls (plasma cortisol) (51), 50 daily smokers and 17 non-daily, and 64 controls (plasma cortisol) where cortisol levels also correlated with craving (52). Two stress-response studies reported negative findings, which employed fear-conditioning (53) and mood induction (54) paradigms to induce stress.

Similar results have been seen using pharmacological stressors such as opioid antagonists. A study employing a pharmacological challenge, using the opioid antagonist naltrexone, in 23 smokers and 25 controls found that both plasma cortisol and ACTH responses were blunted in smokers compared to controls (55). Unsurprisingly, peak ACTH response occurred before peak cortisol response, though salivary cortisol levels were not blunted in the smoking group. A blunted plasma cortisol response to naloxone, another opioid antagonist, was also seen in a study of 9 smokers and 11 controls (56). However, a third naltrexone study did not replicate such findings. Negative results were reported in a study of a dexamethasone challenge to 24 female smokers (57). Overall, these studies indicate that nicotine dependent smokers show blunted physiological responses to stress, across different stress-induction methods, and a potential implication of the opioid system in some of these changes.

Studies of stress response in smokers have also examined effects of acute abstinence on stress responses, where cortisol responses appear to be less robust. A study of 16 non-abstinent, 17 overnight-abstinent (with nicotine patch), and 16 overnight-abstinent (placebo patch) smokers found blunted TSST-induced salivary cortisol responses in the non-abstinent and nicotine patch groups compared to placebo (58). No relationships with subjective effects or symptoms were found. A study of 72 smokers undergoing 24-hour abstinence measured salivary cortisol, and plasma ACTH and cortisol responses to a different stress test, the paced auditory serial addition task (PASAT) (59). The stress protocol increased both ACTH and cortisol, though only ACTH showed a blunted response. A recent study of 20 smokers used a crossover design to determine changes in experimentally-induced craving and blood levels of cortisol, ACTH, and prolactin in response to a 1.6mg or 3.2mg naltrexone challenge (60). Experimental procedures consisted of 4-hour induced abstinence followed by challenge with naltrexone or placebo. Drug administration was followed by smoking-related visual cues or neutral cues. Smoking cues resulted in increased self-reported craving scores. Both naloxone doses resulted in greater levels of cortisol, ACTH, and prolactin in response to smoking-cues, compared to placebo treatment and smoking cues, suggesting a potentiation of the stress response due to cue reactivity during withdrawal. Interestingly, the relationship between cortisol changes and craving may be dissociable, as studied in 37 overnight-abstinent smokers. This study examined the effect of intranasal insulin pre-treatment on blood cortisol levels in response to the TSST (61). Intranasal insulin reduced measures of craving (visual analog scale) and increased pre-TSST cortisol levels which remained elevated after the TSST. Given the potential utility of cortisol responses clinically, it is important to characterize such cases where associations between biomarkers and clinical outcomes are less robust, and the impact of cessation aids.

The impact of longer-term abstinence was assessed in a group of 33 smokers attempting an 8-day period of abstinence, with follow-up assessment 7-days afterwards to assess relapse (62). Plasma cortisol, dehydroepiandrosterone (DHEA – an adrenal-derived steroid hormone), DHEA-sulphate, and cotinine were measured. No individual marker was altered by abstinence at 8-days, though the DHEA:cortisol ratio was lower in those who relapsed at 15 days compared to those who continued abstinence. The DHEA:cortisol ratio has been proposed to more accurately assess net glucocorticoid production given the opposing actions of DHEA and cortisol (63).

### Stress Hormones

In some studies, stress was induced in the form of induced withdrawal, with acute (<24hours) withdrawal being the most common. One such study reported increased salivary cortisol 2 and 3-hours after last cigarette (n=20 male smokers), which was associated with intensity of craving (64). Another study of 60 smokers in 3-hour withdrawal and 64 non-smokers measured plasma orexin, leptin, cortisol and ACTH (65). Though all markers were unchanged from pre-withdrawal levels, craving scores were correlated positively with leptin and negatively with orexin. A study of overnight abstinence in 24 male smokers measured plasma cortisol, ACTH, DHEA, and nicotine acutely after smoking either a regular cigarette or VLNC (66). Interestingly, all hormone levels decreased after VLNC intake but ACTH and DHEA increased after regular cigarette intake. This suggests a further utility of VLNCs in reducing stress responses, which have been implicated in craving in the above studies. Additionally, these findings may be indicative of nicotine-specific effects on these HPA axis biomarkers.

Two studies have examined longer-term withdrawal in the form of cessation trials. A no-treatment cessation trial measured serum cortisol, catecholamines, and prolactin in 33 quit-attempters and 33 non-quit-attempting smokers at intake and after 6-weeks (67). Successful quitters showed decreased cortisol and epinephrine, but no baseline measures alone were predictive of a successful quit attempt. The second study, a 6-week NRT (patch) RCT of 112 smokers, measured salivary cortisol at baseline, 1 day, and 1, 2, and 6-weeks (68). Subjects showed decreased cortisol after 1 day, with successful quitters showing a persistent decrease. Contrary to expectations, greater cortisol decrease after 1 day was associated with greater chance of relapse. These studies show compelling results for the use of cortisol as a predictive marker of treatment response and risk of relapse.

### CVD and Lipid Biomarkers

Smoking is one of the most salient risk factors causally linked to CVD, with a wide body of literature reporting greater levels of CVD risk biomarkers in smoking populations (69, 70). However, a growing body of evidence shows that highly dependent smokers have elevated CVD risk, beyond that which is explained by smoking alone (71). This mirrors findings from psychiatric disorders such as major depressive disorder (MDD) and bipolar disorder (BD) (72) and may be indicative of shared pathophysiological features between nicotine dependence/TUD and CVD.

Commonly reported CVD risk biomarkers include the lipid profile: total cholesterol (TC), high-density lipoprotein cholesterol (HDL), low-density lipoprotein cholesterol (LDL), and triglycerides (TG) of which TC, LDL, and TG are associated with increased CVD risk and HDL is associated with decreased risk. Additional markers measured in the following studies include polyunsaturated fats (PUFAs) which are generally thought to be protective (73) and two calculated risk factors; atherogenic index of plasma (TG:HDL) and the atherogenic coefficient (non-HDL:HDL) (74, 75).

Results from the National Health and Nutrition Survey (NHANES) of 3903 current smokers measuring lipid profile found that an earlier TTFC was associated with lower HDL, greater LDL, and a poorer overall lipid profile (i.e. greater percentage of markers in unhealthy ranges) (76). Other research has found similar results with atherogenic measures, namely a study examining TG, HDL and LDL in 120 patients with TUD alone, 134 patients with comorbid TUD and mood disorders (MDD or BD), and 77 controls (77). Atherogenic index and atherogenic coefficient were both increased in TUD versus controls and were highest in comorbid TUD and mood disorders. Another study, focusing on comorbid disorders, measured plasma malondialdehyde (MDA - a marker of lipid peroxidation and oxidative stress), HDL, and TG in 90 metabolic syndrome patients (48 with TUD) and 224 non-metabolic syndrome controls (82 with TUD) (78). Subjects with TUD showed a greater atherogenic index compared to non-TUD subjects, regardless of metabolic syndrome status. Lastly, a cross-sectional study of 107 male smokers found that serum leptin levels, a hormone which stimulates lipolysis, were negatively correlated with FTND scores, though lipid profile measures did not correlate with any TUD measure (79).

Longitudinal studies have expanded on these cross-sectional findings, indicating potential predictive value of CVD markers. A 90-day RCT of a 3mg daily omega-3 PUFA supplement in 63 smokers measured baseline circulating levels of the omega-3 PUFAs docosahexaenoic acid (DHA) and eicosapentaenoic acid (EPA) (80). A pre-treatment comparison with 51 controls found decreased DHA levels in smokers, and PUFA supplementation resulted in decreased FTND scores at 90 days though no correlational analyses were performed. A 12-week longitudinal study of cessation-induced weight gain measured lipid profile and CRP in 186 treatment-seeking smokers, 89 receiving 21mg nicotine patch and 95 receiving varenicline (81). At baseline and 12-weeks, BMI positively correlated with TG levels and FTND scores, and negatively with HDL. Greater baseline FTND scores and TG levels were predictive of BMI gain at 3 months. BMI changes are notable in this study, as cessation-related weight gain is a well-documented risk factor for medication non-compliance and unsuccessful quit attempts (82). Lastly, a 12-week RCT of exercise therapy assessed lipid profile, red and white blood cell concentration, and C-reactive protein (CRP - a marker of inflammation) in 130 female smokers at baseline and at 12-weeks (83). Those who remained abstinent at 12-weeks had reduced TC and TC/HDL ratio, and greater baseline CRP levels were predictive of relapse at 12-weeks.

Negative findings, or no associations with dependence were performed, in 3 studies. A cross-sectional study of 60 smokers, 48 smokeless tobacco users, and 60 non-tobacco users measuring lipid profile and a panel of hematological markers (84). Another cross-sectional study measuring circulating PUFA levels in 50 smokers and 50 non-smokers (85). Lastly no associations between FTND scores and lipid profile were found in a 12-month RCT of atorvastatin treatment in 160 smokers (86).

Overall, these results suggest that atherogenic markers show more robust associations with severity of dependence and successful treatment responses (i.e. maintenance of abstinence or fewer adverse effects) than individual markers. Though these markers are disrupted in TUD, perhaps suggestive of utility as diagnostic aids, specific associations with measures of dependence or other symptom dimensions are understudied. Thus, the evidence is more tentative for these markers, though they represent an interesting research opportunity given their potential as dual-use biomarkers, for both CVD and TUD outcomes.

### Other Markers

3 studies did not fit into one of the broad categories discussed above. An 8-week study of varenicline as a cessation aid in 20 smokers measured plasma levels of the DA metabolite homovanillic acid (HVA) and the NE metabolite 3-methoxy-4-hydroxyphenylglycol (MHPG) (87). At intake, HVA and MHPG were higher in smokers than a group of 21 controls, suggesting increased baseline monoamine turnover, though neither marker changed after treatment. Unfortunately, two studies did not examine relationships between biomarkers and TUD outcomes. One study of 150 smokers and 191 controls measured a panel of inflammatory and oxidative stress markers (88), and another measuring circulating liver enzymes in 10 controls, 10 patients with colon cancer and combined TUD and alcohol use disorder (AUD), and 8 patients with colon cancer alone (89). However, the presence of severe somatic disease in the second study confounds the biomarker results greatly.

## Discussion

This systematic review yielded many study studies revealing associations between peripheral biomarkers and clinical outcomes, most notably: severity of dependence, treatment response, and risk of relapse after cessation. Biomarkers of exposure such as cotinine, beyond indicating smoking status, showed robust associations with severity of dependence (30-32, 40), and low NMR showed an association with increased risk of relapse during cessation (22, 35). Cotinine showed less consistent associations with CPD, suggesting expired CO as a superior status marker and cotinine/NMR a superior dependence marker. Exposure markers, NMR primarily, currently appear to have the most evidence for their prospective use as treatment response biomarkers (i.e. lower NMR corresponds to greater relapse risk) compared to stress and CVD markers.

Stress marker results suggest they have utility being measured at steady-state levels over time, and in response to a stress challenge. More severely dependent smokers showed more blunted cortisol and ACTH responses (50-52, 55, 59, 68), which may be specific to the presence of nicotine (53, 58), and may indicate a disrupted ability to cope with stressful situations. Cortisol responses to stress may have potential use as a diagnostic aid and longitudinal index of dependence, though this requires a validated and repeatable stress-induction procedure, as no study above performed the TSST more than once. Less consistent results were found with naloxone/naltrexone challenges, with positive and negative findings reported (56, 57). Furthermore, the importance of a standardized stress procedure was highlighted by more robust associations being found in studies using the TSST, versus more negative findings in studies using other psychosocial stressors. Lastly, only one study examined stress hormones as predictive markers of treatment response, though positive findings were observed regarding their prediction of relapse (62). Future studies should examine the utility of these markers prospectively to properly characterize whether baseline stress hormone levels, or stress-induced levels, are predictive of treatment response.

CVD markers showed promising results, though the body of literature was considerably smaller compared to stress or exposure markers. Severity of dependence was associated with less favourable lipid profiles (76), with HDL, LDL, and atherogenicity measures being most robustly associated with a TUD diagnosis (78, 85). TG and PUFA levels showed tentative associations with treatment response, however these were reported in single studies and require replication (80, 81, 86). Tentative evidence was also observed for the diversity of CVD markers measured in other studies (i.e. leptin, CRP, hemodynamic measures) (80, 83, 84). Those markers classified as “others” were underpowered and in highly confounded populations (colon cancer) when positive findings were found (89), or did not include relevant predictive analyses (87).

These findings should be interpreted in the context of the limitations of this review. First, by limiting the search to TUD and nicotine dependence, some relevant studies measuring dependence but not categorized under the MeSH term were inevitability excluded. This was done given the impracticability of including broad terms such as *smoking* (the inclusion of which added nearly 5,000 abstracts), and the desire to not capture an inordinate number of studies not reporting dependence measures. Second, some potential biomarkers may have been missed, as they may not have fallen under the general *biomarker* term in the search. However, given the commonly reported markers in other biomarker reviews (5, 6), this number was likely small. Third, though this systematic review endeavoured to study TUD and dependence in an objective manner, the information provided by the 6-item FTND, the primary dependence scale, limits the aspects of dependence able to be studied. Further development of self-report and interview-based tools for assessing dependence is warranted.

Overall, these findings indicate that cotinine, NMR, and stress markers (cortisol and ACTH) emerge as lead biomarkers of dependence and treatment response, and prospective studies of their clinical utility are warranted. Longitudinal studies of other biomarkers, particularly CVD markers, presented here should be performed, given the preponderance of cross-sectional study designs and their inability to determine directionality of associations. Studies integrating genetics and neuroimaging markers with peripheral biomarkers were not found in the literature search and would improve the understanding of how these peripheral biomarkers relate to and interact with genetic predispositions and the underlying neurobiology of TUD. Importantly, the absence of a mechanistic understanding of how a peripheral biomarker relates to the CNS need not prevent its use clinically, if it shows a robust correlation with clinically relevant measures.

Finally, this review found that composite biomarkers (e.g. NMR, atherogenic indices) showed, generally, more robust results and greater predictive power than single biomarkers. Future studies should further this trend and examine novel composite biomarkers through statistical methods (e.g. propensity scoring, ROC, principal component analyses, or machine learning approaches) and *a priori* knowledge that best predict symptom severity, treatment response, and risk of relapse. Such studies will improve the treatment of TUD and may allow personalized prognoses and therapies based on biomarker profiles.

## Methods

A systematic review of the literature on peripheral biomarkers in the context of TUD was performed using the Preferred Reporting Items for Systematic Reviews and Meta-Analyses (PRISMA) statement (27). MEDLINE searches were conducted of all articles from 1946 to April week 2, 2019 using the following medical subject heading (MeSH) terms: *biomarkers, dihydroxyacetone phosphate, bilirubin, inositol, cotinine, adrenocorticotropic hormone, cortisol, pituitary-adrenal system, homovanillic acid, dopamine, pro-opiomelanocortin, lipids, lipid metabolism* each cross-referenced with *tobacco-use disorder*. These search terms were selected based on established TUD pathophysiology (DA and nicotine metabolism) and lead biomarkers presented in a recent non-systematic review (6). The resulting search was limited to human studies, and abstracts were screened using the inclusion and exclusion criteria described below, included studies were read in full. *Nicotine dependence* was not included as an independent MeSH term as it is a nested term within *tobacco use disorder*.

Studies were included if they reported quantitative measures of peripheral biological compounds in individuals diagnosed with TUD, objectively measured as nicotine dependent, or undergoing cessation treatment. Studies without human subjects, those with SUD populations that did not report TUD findings independently, or those without original research findings (e.g. reviews or meta-analyses) were excluded. Studies whose biological markers were solely genetic were also excluded, given the focus of this review on dynamic biomarkers. Studies measuring only expired CO were not included if this measure was used solely for assessing smoking status.

## Data Availability

Not applicable, review article

